# The Spike-specific IgA in milk commonly-elicited after SARS-Cov-2 infection is concurrent with a robust secretory antibody response, exhibits neutralization potency strongly correlated with IgA binding, and is highly durable over time

**DOI:** 10.1101/2021.03.16.21253731

**Authors:** Alisa Fox, Jessica Marino, Fatima Amanat, Kasopefoluwa Oguntuyo, Jennifer Hahn-Holbrook, Benhur Lee, Florian Krammer, Susan Zolla-Pazner, Rebecca L. Powell

## Abstract

Approximately 10% of infants will experience COVID-19 illness requiring advanced care (1). A potential mechanism to protect this population could be provided by passive immunity through the milk of a previously infected mother. We and others have reported on the presence of SARS-CoV-2-specific antibodies in human milk (2-5). We now report the prevalence of SARS-CoV-2 IgA in the milk of 75 COVID-19-recovered participants, and find that 88% of samples are positive for Spike-specific IgA. In a subset of these samples, 95% exhibited robust IgA activity as determined by endpoint binding titer, with 50% considered high-titer. These IgA positive specimens were also positive for Spike-specific antibodies bearing the secretory component. Levels of IgA antibodies and antibodies bearing secretory component were shown to be strongly positively correlated. The secretory IgA response was dominant among the milk samples tested compared to the IgG response, which was present in 75% of samples and found to be of high-titer in only 13% of cases. Our IgA durability analysis using 28 paired samples, obtained 4-6 weeks and 4-10 months after infection, found that all samples exhibited persistently significant Spike-specific IgA, with 43% of donors exhibiting increasing IgA titers over time. Finally, COVID-19 and pre-pandemic control milk samples were tested for the presence of neutralizing antibodies; 6 of 8 COVID-19 samples exhibited neutralization of Spike-pseudotyped VSV (IC_50_ range, 2.39 – 89.4ug/mL) compared to 1 of 8 controls. IgA binding and neutralization capacities were found to be strongly positively correlated. These data are highly relevant to public health, not only in terms of the protective capacity of these antibodies for breastfed infants, but also for the potential use of such antibodies as a COVID-19 therapeutic, given that secretory IgA is highly stable not only in milk and the infant mouth and gut, but in all mucosa including the gastrointestinal tract, upper airway, and lungs (6).

## Background

Though COVID-19 pathology among children is typically more mild compared to adults, approximately 10% of infants under the age of one year experience severe COVID-19 illness requiring advanced care, and an ever-growing number of children appear to exhibit signs of “Multisystem Inflammatory Syndrome in Children (MIS-C) associated with COVID-19” weeks or months after exposure (1, 7). Furthermore, infants and young children can also transmit SARS-CoV-2 to others and the efficacy of vaccines available for adults have not yet been evaluated for young children or infants (8). Certainly, protecting this population from infection is essential (9).

One potential mechanism of protection is passive immunity provided through breastfeeding by a previously-infected mother. Mature human milk contains ∼0.6mg/mL of total immunoglobulin (10). Approximately 90% of human milk antibody (Ab) is IgA, nearly all in secretory (s) form (sIgA, which consists of polymeric Abs complexed to J-chain and secretory component (SC) proteins) (11). Nearly all sIgA derives from the gut-associated lymphoid tissue (GALT), via the *entero-mammary link*, though there is also homing of B cells from other mucosa (e.g., from the respiratory system), and possibly drainage from local lymphatics of systemic IgA to the mammary gland (11). Unlike the Abs found in serum, sIgA found in milk is highly stable and resistant to enzymatic degradation not only in milk and the infant mouth and gut, but in all mucosae including the gastrointestinal tract, upper airway, and lungs (6). Notably, after two hours in the infant stomach, the total IgA concentration decreases by <50%, while IgG concentration decreases by >75%; importantly, though total SC concentration decreases by ∼60%, there ia no decrease in the stomach of infants born pre-term (within the first 3 months of life) – a population highly vulnerable to infection (12).

Previously we reported on 15 milk samples obtained early in the pandemic from donors recently-recovered from a confirmed or suspected case of COVID-19 (2). In that preliminary study, it was found that all samples exhibited significant IgA binding activity against the SARS-CoV-2 Spike. Eighty percent of samples further tested for Ab binding reactivity to the receptor binding domain (RBD) of the Spike exhibited significant IgA binding, and all of these samples were also positive for RBD-specific secretory Ab reactivity with only small subsets of samples exhibiting specific IgG and/or IgM activity, strongly suggesting the RBD-specific IgA was sIgA. In the present study, we report on the prevalence and isotypes of Spike-specific milk Ab from a larger cohort of donors obtained 4-6 weeks post-confirmed SARS-CoV-2 infection, on the durability of these Abs up to 10 months post-infection, and on SARS-CoV-2-directed neutralization by Abs in a subset of these samples,

## Methods

### Study participants

Individuals were eligible to have their milk samples included in this analysis if they were lactating and had a laboratory-confirmed SARS-CoV-2 infection 4-6 weeks prior to the initial milk sample used for analysis. Certain participants were also able to continue participation in the study and provide a follow-up sample 4-10 months after confirmed infection. This study was approved by the Institutional Review Board (IRB) at Mount Sinai Hospital (IRB 19-01243).

Milk was frozen in participants’ home freezers until samples were picked up and stored at −80°C until Ab testing. Pre-pandemic negative control milk samples were obtained in accordance with IRB-approved protocols prior to December, 2019 for other studies, and had been stored in laboratory freezers at −80°C before processing following the same protocol described for COVID-19 milk samples.

### ELISA

Levels of SARS-CoV-2 Abs in human milk were measured as previously described (2). Briefly, before Ab testing, milk samples were thawed, centrifuged at 800g for 15 min at room temperature, fat was removed, and the de-fatted milk transferred to a new tube. Centrifugation was repeated 2x to ensure removal of all cells and fat. Skimmed acellular milk was aliquoted and frozen at −80°C until testing. Both COVID-19 recovered and control milk samples were then tested in separate assays measuring IgA, IgG, and secretory-type Abs, in which the secondary Ab used for the latter measurement was specific for free and bound SC. Half-area 96-well plates were coated with the full trimeric recombinant Spike protein produced, as described previously (13). Plates were incubated at 4°C overnight, washed in 0.1% Tween 20/PBS (PBS-T), and blocked in PBS-T/3% goat serum/0.5% milk powder for 1 h at room temperature. Milk was used undiluted or titrated 4-fold in 1% bovine serum albumin (BSA)/PBS and added to the plate. After 2h incubation at room temperature, plates were washed and incubated for 1h at room temperature with horseradish peroxidase-conjugated goat anti-human-IgA, goat anti-human-IgG (Fisher), or goat anti-human-secretory component (MuBio) diluted in 1% BSA/PBS. Plates were developed with 3,3’,5,5’-Tetramethylbenzidine (TMB) reagent followed by 2N hydrochloric acid (HCl) and read at 450nm on a BioTek Powerwave HT plate reader. Assays were performed in duplicate and repeated 2x.

### IgA extraction from milk

Total IgA was extracted from 25 – 100mL of milk using peptide M agarose beads (Pierce) following manufacturer’s protocol, concentrated using Amicon Ultra centrifugal filters (10 kDa cutoff; Millipore Sigma) and quantified by Nanodrop.

### Pseudovirus neutralization assay

Neutralization assays were performed using a standardized SARS-CoV-2 Spike-pseudotyped Vesicular Stomatitis Virus (VSV)-based assay with ACE2- and TMPRSS2-expressing 293T cells as previously described (14). Briefly, pseudovirus was produced by transfection of 293T cells with SARS-CoV-2 Spike plasmid, followed 8 h later by infection with a VSVΔG-rLuc reporter virus. Two days post-infection, supernatants were collected and clarified by centrifugation (14). A pre-titrated amount of pseudovirus was incubated with serial dilutions of extracted IgA for 30 min at room temperature prior to infection of cells seeded the previous day. Twenty hours post-infection, cells were processed and assessed for luciferase activity as described (14).

### Analytical Methods

Control milk samples obtained prior to December 2019 were used to establish positive cutoff values for each assay. Milk was defined as positive for the SARS-CoV-2 Abs if OD values measured using undiluted milk from COVID-19-recovered donors were two standard deviations (SD) above the mean ODs obtained from control samples. Endpoint dilution titers were determined from log-transformed titration curves using 4-parameter non-linear regression and an OD cutoff value of 1.0. Endpoint dilution positive cutoff values were determined as above. Percent neutralization was calculated as (1- (average luciferase RLU of triplicate test wells – average luciferase expression RLU of 6 ‘virus only’ control wells) *100. Mann-Whitney U tests were used to assess significant differences in the grouped COVID-19-recovered and control extracted IgA neutralization capacities. The concentration of milk IgA required to achieve 50% neutralization (IC_50_) was determined as described above for endpoint determination. Correlation analyses were performed using Spearman correlations. All statistical tests were performed in GraphPad Prism, were 2-tailed, and significance level was set at p-values < 0.05.

## Results

### Ab profile in milk from COVID-19-recovered donors 4-6 weeks after infection

Skimmed acellular milk was aliquoted and frozen at −80° C until testing. Undiluted milk samples obtained 4-6 weeks post-infection from 75 COVID-19-recovered donors, and 20 pre-pandemic milk samples obtained prior to December, 2019 were screened in our IgA ELISA against recombinant trimeric SARS-CoV-2 Spike. Sixty-six of 75 samples (88%) were positive for Spike-specific IgA, with the COVID-19 samples exhibiting significantly higher Spike-specific IgA binding compared to controls (**Fig. 1a;** p<0.0001). Following this initial screening, 40 of the Spike-positive samples were further titrated to determine binding endpoint titers as an assessment of Ab affinity and/or quantity (**Fig. 1b**). Thirty-eight of 40 (95%) Spike-reactive samples exhibited positive IgA endpoint titers and 19 of these samples (50%) were ≥5 times higher than the endpoint titer of the positive cutoff value, and were therefore designated as ‘high-titer’ (**Fig.1c**).

**Figure 1.**
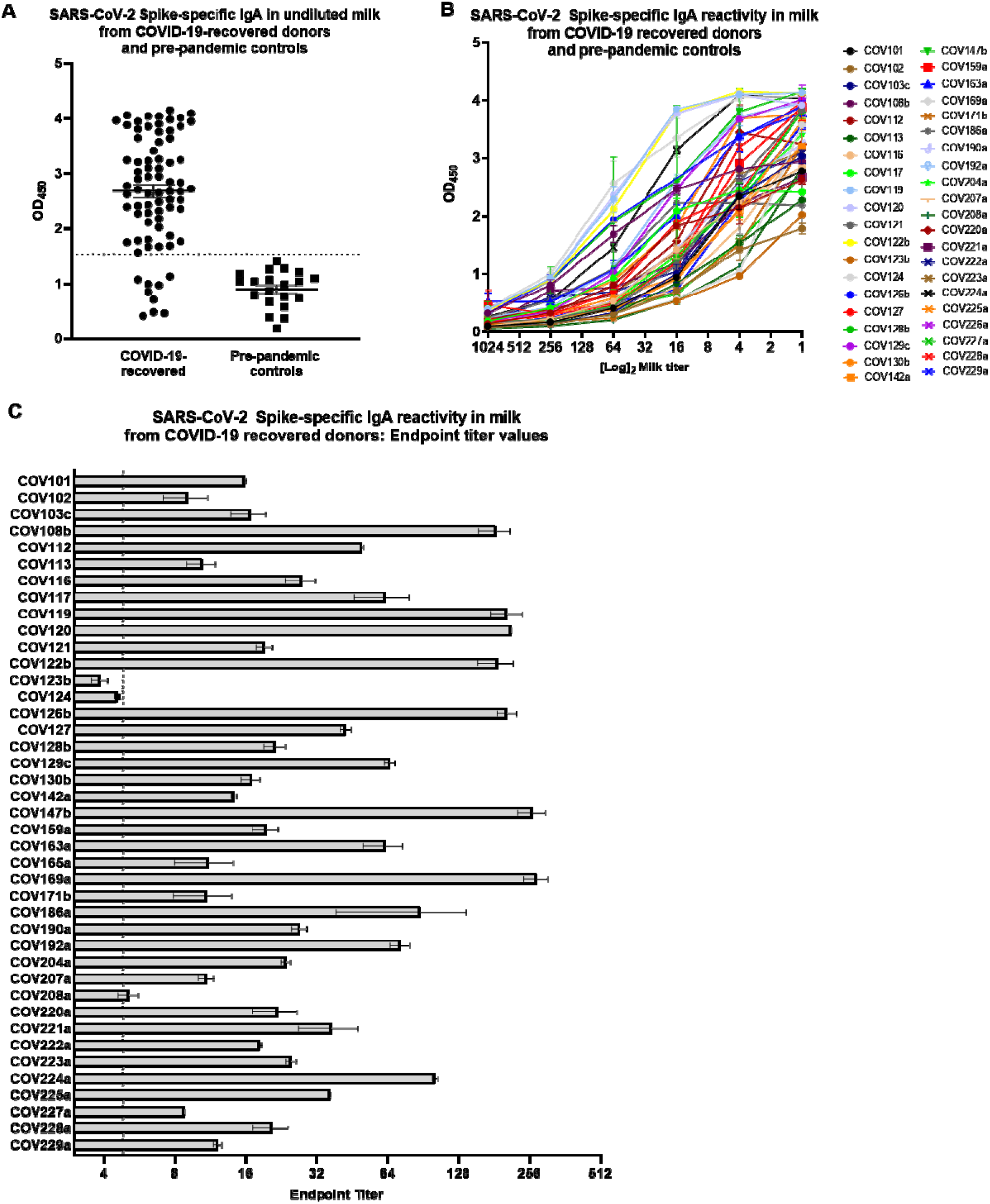
A robust, Spike-specific IgA response in milk commonly occurs after SARS-CoV-2 infection. (A) Screening of undiluted milk samples for specific IgA by ELISA against the full-length Spike trimer. Mean values with SEM are shown. Dotted line: positive cutoff value (mean OD of negative control milk samples + 2*SD). (B) Full titration against Spike of 40 milk samples found to be positive by the initial screening. (C) Endpoint dilution titers of the 40 titrated milk samples. Segmented line: positive cutoff value; dotted line: 5x positive endpoint cutoff value, designating samples as ‘high-titer’.

Additionally, 20 samples assayed for Spike-specific IgA were also assessed for Spike-specific secretory Ab (by detecting for SC), and IgG. Nineteen of these undiluted milk specimens (95%) from convalescent COVID-19 donors were positive for Spike-specific secretory Abs compared to pre-pandemic control milk (**Fig. 2a**). One sample (COV125) was negative for specific IgA but positive for specific secretory Ab, while another sample (COV123) was positive for specific IgA but negative for specific secretory Ab. Eighteen undiluted milk samples (95%) exhibiting Spike-specific secretory Ab activity also exhibited positive endpoint titers (**Fig. 2c**). Of the samples found to be high-titer for Spike-specific IgA, 7 were also high-titer for specific secretory Ab (70%). Mean OD values for undiluted milk and endpoint titers were used in separate Spearman correlation tests to compare IgA and secretory Ab reactivity (**Fig. 2e**). It was found that IgA and secretory Ab levels were positively correlated (using ODs: r=0.77, p<0.0001; using endpoint titers: r=0.86, p<0.0001). Additionally, 15/20 undiluted milk samples from COVID-19-recovered donors were positive for Spike-specific IgG compared to pre-pandemic controls (75%; **Fig. 2b**), with 13/15 of these samples exhibiting a positive endpoint titer (87%; **Fig. 2d**), and 2/15 designated as high titer with values ≥5 times cutoff (13%). No correlation was found between IgG and IgA titers or between IgG and SC titers (data not shown).

**Figure 2.**
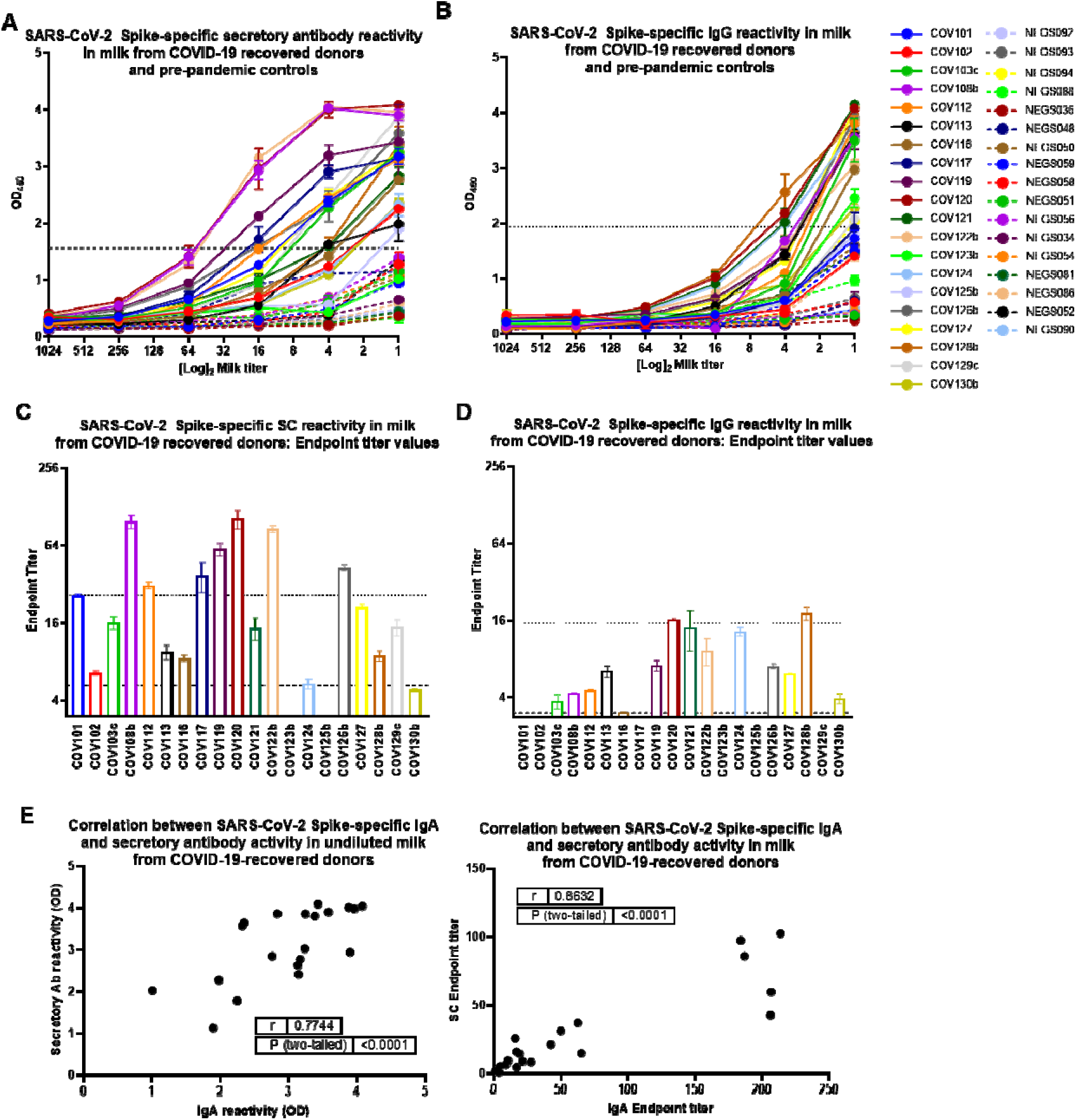
The dominant Spike-specific IgA response in milk after SARS-CoV-2 infection is strongly correlated with a robust secretory Ab response, while specific IgG activity is relatively modest. Twenty samples assayed for Spike-specific IgA were also assessed for Spike-specific secretory Ab (by detecting for SC), and IgG. (A, B) Full titration against Spike, detecting (A) secretory Ab, and (B) IgG. NEG (i.e. negative)/segmented lines: pre-pandemic controls. COV/solid lines: milk from COVID-19-recovered donors. Dotted lines: positive cutoff values. (C, D) Endpoint titer values calculated for (A) secretory Ab, and (B) IgG. Segmented lines: positive cutoff values; dotted lines: 5x positive cutoff (high-titer cutoff). (E) IgA and secretory Ab binding OD values or endpoint titers were used in 2-tailed Spearman correlation tests. SC: secretory component.

### Durability of the SARS-CoV-2 Spike-specific milk IgA response

To assess the durability of this sIgA-dominant response, 28 pairs of milk samples obtained from COVID-19-recovered donors 4-6 weeks and 4-10 months after infection were assessed for Spike-specific IgA. All donors exhibited persistently significant Spike-specific IgA titers of the period of follow-up. Mean endpoint titers from the early to the late milk samples grouped were not significantly different (Fig. 3a). Fourteen donors (50%) exhibited >10% decrease in IgA titer, 12 donors (43%) exhibited >10% increase in IgA titer, and 2 donors (7%) exhibited no change in titer (**Fig. 3a**). Notably, only 2 donors (7%) exhibited >50% decrease in titer over time. Furthermore, examining a subset of these samples with the longest follow-up, obtained 7-10 months after infection, mean endpoint titers measured from the early to the late milk samples were also not significantly different (data not shown). These longest follow-up samples included 5 donors (36%) with >10% decrease in IgA titer, 8 donors (57%) with >10% increase in IgA titer, and 1 donor (7%) with no change in titer (**Fig. 3b**). Only 1 donor (7%) exhibited >50% decrease in titer, and as with the larger durability cohort, all donors exhibited persistently significant Spike-specific IgA titers.

**Figure 3.**
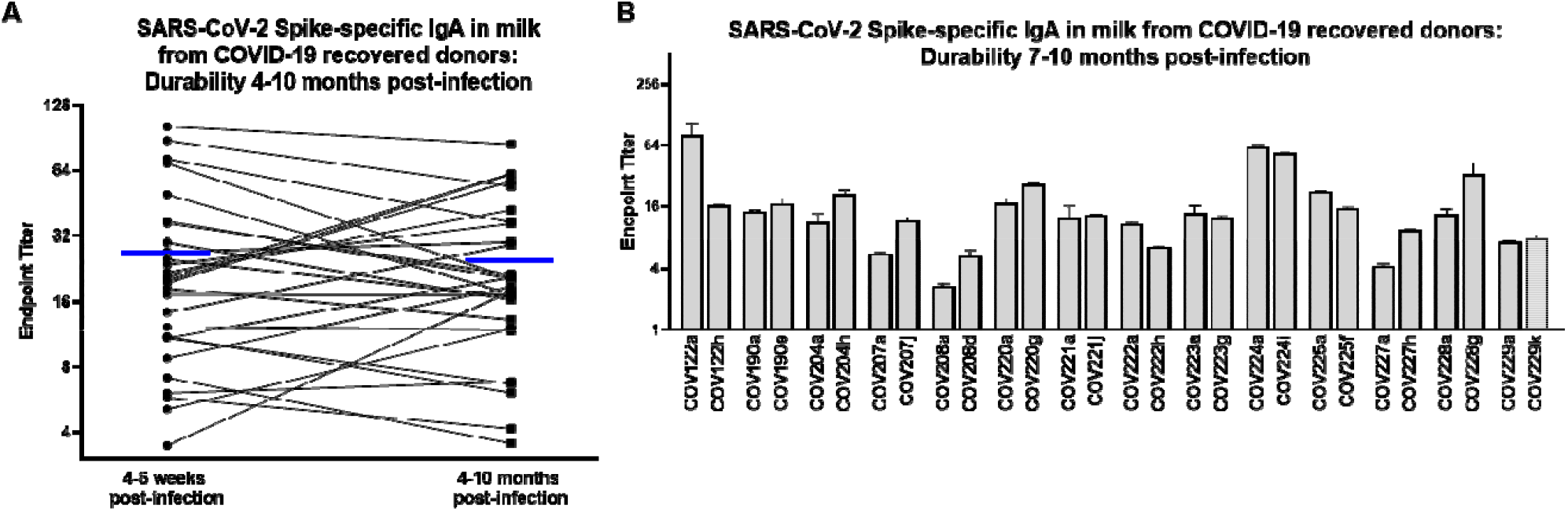
The Spike-specific IgA response in milk after SARS-CoV-2 infection is highly durable over time. (A) IgA endpoint titers determined from Spike ELISA for 28 pairs of milk samples obtained from COVID-19-recovered donors 4-6 weeks and 4-10 months after infection are shown. Blue lines indicate mean endpoint values for each group. (B) IgA endpoint titers for a subset of 14 paired samples obtained 4-6 weeks and 7-10 months after infection. Mean with SEM is shown.

### SARS-CoV-2 neutralization capacity of total milk IgA from COVID-19-recovered donors

Total IgA was extracted from 8 COVID-19 and 8 control milk samples analyzed for Spike-specific Ab profile. All 8 COVID-19 samples had been shown to exhibit positive Spike-specific IgA and secretory Ab titers (**Figs 1-2**). Neutralization capacity was tested using a Vesicular Stomatitis Virus (VSV)-based pseudovirus assay, wherein the native VSV surface protein G is replaced by the SARS-CoV-2 Spike, as described previously ((14); **Fig. 4**). At the maximum concentration tested (200ug/mL total purified milk IgA), 6/8 (75%) COVID-19 samples exhibited >50% neutralization (mean, 87% neutralization; range, 70% - 100%), while only 1/8 control samples (13%) achieved this benchmark (94% neutralization; **Fig. 4a**). Mean percent neutralization values at 50ug/ml extracted IgA were grouped and compared among COVID-19 and pre-pandemic control samples. COVID-19 samples exhibited significantly greater neutralization compared to controls (p=0.0064; **Fig. 4b**). As well, when the concentration of IgA required to achieve 50% neutralization (IC50) was determined, 7/8 pre-pandemic controls did not achieve 50% neutralization (IC50>200ug/mL while, for the COVID-19 samples, 2/8 did not achieve 50% neutralization, and the mean IC50 for the 6 COVID19 specimens that displayed neutralizing activity was 33.6ug/mL of total IgA (range, 2.39 – 89.4ug/mL; **Fig. 4c**). Finally, we compared the neutralization IC50 titers to the IgA endpoint titers measured for these samples (**Fig. 1**). There was a significant positive correlation between IgA binding and neutralization capacities (r=0.83, p=0.0154; **Fig. 4d**). Notably, the 2 non-neutralizing COVID-19 IgA samples also exhibited the lowest IgA endpoint titers (COV121, COV130; mean IgA endpoint titers of 19 and 17, respectively), while the 6 neutralizing samples exhibited high Spike-specific IgA binding titers (**Figs. 1c, 4c**).

**Figure 4.**
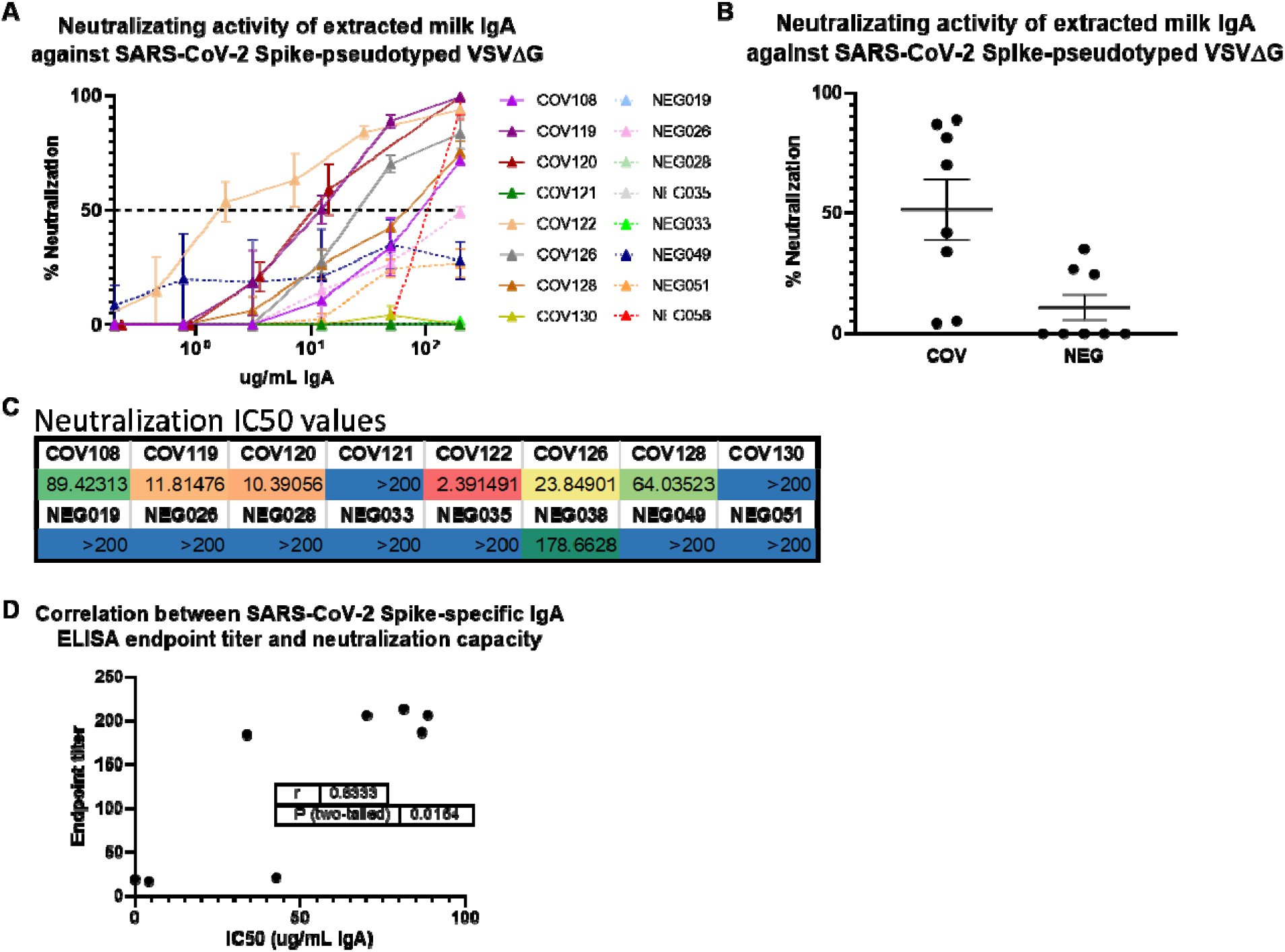
Extracted milk IgA from COVID-19-recovered donors exhibits SARS-CoV-2 Spike-targeted neutralization potency that is highly correlated with IgA binding activity. (A) Total IgA was purified from milk by conventional means using peptide M agarose. IgA was titrated and tested in a VSV-based SARS-CoV-2 pseudovirus neutralization assay. NEG/segmented lines: pre-pandemic controls. COV/solid lines: COVID-19-recovered milk samples. Segmented line: 50% neutralization cutoff value. (B) Percent neutralization achieved using 50ug/mL of total extracted milk IgA. Mean values with SEM are shown. (C) Neutralization IC50 values determined from IgA titration curves. (D) Endpoint titer values determined in Fig. 1 and IC50 values were used in a 2-tailed Spearman correlation test.

## Discussion

There has been no evidence that SARS-CoV-2 transmits via human milk, with sporadic cases of viral RNA (not infectious particles) detected on breast skin (3); however, there have been reports of viral RNA in the milk (reviewed in (15)), though collection methods in these reports did not necessarily include masking, cleaning of the breast, or even handwashing to avoid contamination from the donor’s environment. As such, the WHO and CDC recommend that infants not be separated from SARS-CoV-2-infected mothers, and that breastfeeding should be established and not disrupted (depending on the mothers’ desire to do so), in combination with masking and other hygiene efforts (16, 17).

We and others have reported SARS-CoV-2-specific Abs in milk obtained from donors with previously confirmed or suspected infection (2-5). Here, we have significantly expanded our earlier work in which we reported on SARS-CoV-2 Ab prevalence among 75 COVID-19-recovered participants whose milk samples were obtained 4-6 weeks after confirmed SARS-CoV-2 infection. Indeed, we have confirmed among this much larger sample set that a SARS-CoV-2 IgA Ab response in milk after infection is very common. This IgA response dominates compared to the measurable but relatively less frequent IgG response that is markedly less robust. Importantly, a very strong positive correlation was found between Spike-specific milk IgA and secretory Abs, using both ELISA OD values of Ab binding in undiluted milk as well as Ab binding endpoint titers, indicating that a very high proportion of the SARS-CoV-2 Spike-specific IgA measured in milk after SARS-CoV-2 infection is sIgA, confirming our early reports. This is relevant for the effective protection of a breastfeeding infant, given the high durability of secretory Abs in the relatively harsh mucosal environments of the infant mouth and gut (6, 12). These data are also relevant to the possibility of using extracted milk IgA as a COVID-19 therapy. Extracted milk sIgA used therapeutically would likely survive well upon targeted respiratory administration, with a much lower dose of Ab likely needed for efficacy compared to systemically-administered convalescent plasma or purified plasma immunoglobulin.

All COVID-19 IgA samples analyzed that had been designated as ‘high titer’ for Spike-specific IgA exhibited significant Spike-directed neutralization capacity, wherein IgA binding endpoint titers and neutralization IC50 values were found to be significantly correlated. Of the 3 samples examined for neutralization capacity that exhibited positive but not high titer Spike-specific IgA, 2 were non-neutralizing. It should be noted that these were all samples obtained 4-6 weeks after infection, and future samples may exhibit neutralization as the Ab response matures. These data extend the recent analyses of SARS-CoV-2 neutralization using diluted whole milk (3, 5).

Critically, our IgA durability analysis using 28 paired samples obtained 4-6 weeks and 4-10 months after infection revealed that for all donors, Spike-specific IgA titers persisted for as long as 10 months, a finding that is highly relevant for protection of the breastfeeding infant over the course of lactation, and also pertinent to the size of a potential donor pool for collection of milk from COVID-19-recovered donors for therapeutic use of extracted milk IgA. Notably, even after 7-10 months, only 5 of 14 samples exhibited >10% decrease in specific IgA endpoint titers, while 8 of 14 samples actually exhibited an increase in specific IgA titer. These highly durable or even increased titers may be reflective of long-lived plasma cells in the GALT and/or mammary gland, as well as continued antigen stimulation in these compartments, possibly by other human coronaviruses, or repeated exposures to SARS-CoV-2.

Given the present lack of knowledge concerning the potency, function, durability, and variation of the human milk immune response not only to SARS-CoV-2 infection, but across this understudied field in general, the present data contributes greatly to filling immense knowledge gaps and furthers our work towards *in vivo* efficacy testing of extracted milk Ab in the COVID-19 pandemic context and beyond.

## Data Availability

The authors confirm that the data supporting the findings of this study are available within the article.

## Acknowledgements

As always, we are indebted to the milk donors who make this work possible. This work was supported by the NIH/NIAID.

